# Evidence of reduced inhibition in older adults with a history of repetitive brain trauma. A transcranial magnetic stimulation study

**DOI:** 10.1101/2023.03.18.23287431

**Authors:** Alan J Pearce, Dawson J Kidgell, Ashlyn K Fraser, Billymo Rist, Jamie Tallent

## Abstract

International concern regarding the association between repetitive neurotrauma in sport and long term concerns with ageing continues. While previous studies have reported older (i.e. over 50 years) our study describes corticomotor changes across the lifespan between retired contact sport athletes, between the ages of 30 and 70 years. Retired athletes, minimum five years retired, (n=152; 48.6±9.0 years) and age-matched controls (n=72; 47.8±9.5 years) were assessed using single and paired-pulse transcranial magnetic stimulation (TMS) for active motor threshold (aMT), motor evoked potential and cortical silent period duration (expressed as MEP:cSP ratio), and short- and long-interval intracortical inhibition (SICI and LICI). Age-matched controls showed significant moderate correlations for MEP:cSP ratios at 130% (*rho*=0.48), 150% (*rho*=0.49)and 170% aMT (*rho*=0.42; all *p*<0.001) and significant but small negative correlation for SICI (*rho*=-0.27; *p*=0.030), and moderate negative correlation for LICI (*rho*=-0.43; *p*<0.001). Further, group-wise correlation analysis shows significant stronger corelations (all *p*<0.05) in the control for each variable than in the retired players. This study is the first to characterise corticomotor differences between retired athletes and age matched controls across the lifespan. in those with a history of repetitive head trauma and provides a foundation for further work to utilise TMS as a prodromal marker useful in supplementing neuropsychological assessment for traumatic encephalopathy syndrome which currently lacks physiological biomarkers.

## Introduction

Concerns regarding sports and the long-term effects of repetitive head impacts on neurological function and risk of neurodegenerative disease continues. Epidemiological studies report, in those with a history of repetitive neurotrauma from playing contact sports, an increased risk of neurological impairments and neurodegenerative disease [1, 2]. While definitive evidence of neurodgenerative disease only exsists post-mortem [3, 4], some attempt has been made to investigate pathophysiological changes in retired contact sport athletes using transcranial magnetic stimulation (TMS) [5-9]. However these studies have been limited to older retired players (<50 years and older) to the exclusion of younger retired players aged in their 30s and 40s. To date, no study has explored TMS changes across the lifespan in this cohort.

Research in the neurophysiology of healthy ageing [10] have utilised single-pulse TMS measures (motor evoked potential [MEP] and cortical silent period [cSP]), as well as paired-pulse TMS short-interval intracortical inhibition (SICI) and long-interval intracortical inhibition (LICI). Both single and paired pulse TMS have been used in cross-sectional studies quantifying intracortical physiology of older retired athletes compared to age-matched controls reporting changes in intracortical inhibition [5-8]. However, with studies focussed on older retired contact sport athletes, no study has yet characterised changes in evoked potentials across the lifespan to include younger retired contact sport athletes with a history of repetitive brain trauma. This descriptive study presents data comparing TMS single and paired pulse measures in retired contact sport athletes, compared to similar age-ranged control cohort of individuals who have never participated in contact sport.

## Methods

### Participants

Retired contact sport athletes (n=152) and age-matched controls (n=72) ranging between 30 and 70 years were recruited for this study. Retired athletes were recruited from a number of different sports including Rugby union and league (n=35), Australian football (n=78), boxing (n=2), motor racing (n=2) and cycling (BMX jumping, n=13). Age-matched controls were recruited from the community following word of mouth and recruitment advertisements. Table 1 outlines all participant details.

**Table 1.** Participant characteristics. Mean (± SD).

|  | **Age**<br>**(years)** | **Height**<br>**(cm)** | **Weight**<br>**(kg)** | **Time since**<br>**retirement (years)** |
| --- | --- | --- | --- | --- |
| Retired<br>athletes<br>(n=152) | 48.6 (±9.0) | 183.4 (±10.5) | 92.8 (±15.9) | 18.5 (±8.9) |
| Controls<br>(n=72) | 47.8 (±9.5) | 179.7 (±7.6) | 89.3 (±12.2) | n/a |

Individuals participating in the study did not report any neurological condition, specific sleep disorders (e.g., obstructive sleep apnoea), or psychiatric disorders, nor any brain injury outside of contact sports participation (e.g., car or workplace accidents, fights or falls). Participants provided voluntary written informed consent prior to data collection. Ethical approval for all protocols were given by the La Trobe University Human Research Ethics Committee (Human Ethics Committee approval number 18005) conforming to the guidelines set out by the Declaration of Helsinki.

### Transcranial magnetic stimulation and electromyography recording

Applying previously described TMS protocols [11-13], TMS was applied over the contralateral primary motor cortex with surface electromyography (sEMG) recording 500 ms sweeps (100 ms pre-trigger, 400 ms post-trigger; PowerLab 4/35, ADInstruments, Australia). The sEMG activity was recorded using bipolar Ag/AgCl electrodes, with an intra-electrode distance of 2 cm positioned over the first dorsal interosseous (FDI) muscle of the participant’s dominant hand adhering to the Non-Invasive Assessment of Muscles (SENIAM) guidelines for sEMG [14].

TMS was delivered using a MagStim 200^*2*^ stimulator (Magstim, UK) using a figure of eight D70 remote coil (Magstim, UK). For reliability of coil placement participants wore a snugly fitted cap (EasyCap, Germany), positioned with reference to the nasion-inion and interaural lines. The cap was marked with sites at 1 cm spacing in a latitude-longitude matrix to ensure reliable coil position throughout the testing protocol [15].

Following identification of the ‘optimal site’, defined as the site with the largest observed MEP [15], active motor threshold (aMT) was determined, during a controlled, low-level voluntary contraction of the FDI muscle at 10% of Maximal Voluntary Contraction (MVC). The aMT was identified by delivering TMS stimuli (5% of stimulator output steps, and in 1% steps closer to threshold) at intensities from a level below the participant’s threshold until an observable MEP of at 200 *µ*V and associated cSP could be measured in at least five of ten stimuli [16]. Stimuli were delivered in random intervals (between 6–10 s) at intensities of 130%, 150% and 170% of aMT. Twenty stimuli were presented in random intervals of four sets of five pulses per set, with a break of 30 s provided between sets and intensity levels to reduce the possibility of muscular fatigue [13].

MEP amplitudes were measured from the peak-to-trough difference of the waveform. Duration of the cSP was calculated from the onset (deflection) of the MEP waveform to the return of uninterrupted EMG [17].

Paired-pulse MEPs for short latency intracortical inhibition (SICI) and long intracortical inhibition (LICI) were measured with the FDI using an interstimulus interval (ISI) of 3 ms and 100 ms respectively. SICI was undertaken with a conditioning stimulus of 80% aMT and a test stimulus of 130% aMT, while LICI was completed using a suprathreshold conditioning and test stimulus at 130% aMT; stimulation ratios and ISIs which we have previously published in post-concussion population groups [8, 18]. Twenty sweeps were delivered, in four sets of five, at random intervals between 8–10 s between stimuli and 30 s between sets to reduce fatigue.

### Data and statistical analyses

Single pulse MEPs were expressed as a cSP:MEP ratio to reflect a balance between excitatory and inhibitory mechanisms, previously shown to reduce between-participant variability [19, 20]. SICI was calculated as a ratio of the paired-pulse MEP at 3ms to the single pulse MEP at 130% aMT. LICI was calculated as a ratio of the suprathreshold test stimulus (1 mV) to the conditioning stimulus [21].

Data was analysed using Jamovi software (Version 2.3, Sydney, Australia). Age, height, and weight were compared using independent *t*-tests. As TMS data was found to be non-normally distributed, relationships between TMS variables and age were explored using Spearman’s *rho*. Interpretations of correlations were used following the following descriptive criteria: <0.1= *trivial*, 0.1 = *small*, 0.3 = *moderate*, 0.5 = *large*, 0.7 = *very large*, 0.9 = *nearly perfect*, 1 = *perfect* [22]. Data is presented as means (±SD) and alpha was set at 0.05. Comparison of correlations between groups was completed using the method described by Cohen et al [23].

## Results

No differences were observed between groups for age (*t*_200_=0.997, *p*=0.319), height (*t*_200_=1.856, *p*=0.074) and weight (*t*_200_=1.621, *p*=0.106). Active motor threshold (aMT) and single pulse cSP:MEP ratios for stimulus intensities 130% aMT, 150% aMT and 170% aMT are presented in figures 1 and 2 respectively. No correlations were found between age and aMT in both groups (fig 1a and b). Correlations between cSP:MEP ratios and age showed large positive significant correlations for the control group for 130% aMT (*rho*=0.503; *p*<0.001, fig 2a), 150% aMT (*rho*=0.513; *p*<0.001, fig 2c) and 170% aMT (*rho*=0.513; *p*<0.001, fig 2e). Conversely, no significant correlations were found for the retired athlete group at each stimulus intensity (fig 2b, 2d, 2f respectively). Comparisons between groups showed no significant difference in correlation for aMT (*Z*=0.328, *p*=0.743). Comparison between groups showed significant correlations in the control group compared to the retired athlete group for cSP:MEP ratios between groups for 130% aMT (*Z*=2.547, *p*=0.011), 150% aMT (*Z*=2.245, *p*=0.026), and 170% aMT (*Z*=1.974, *p*=0.048).

**Figure 1.**
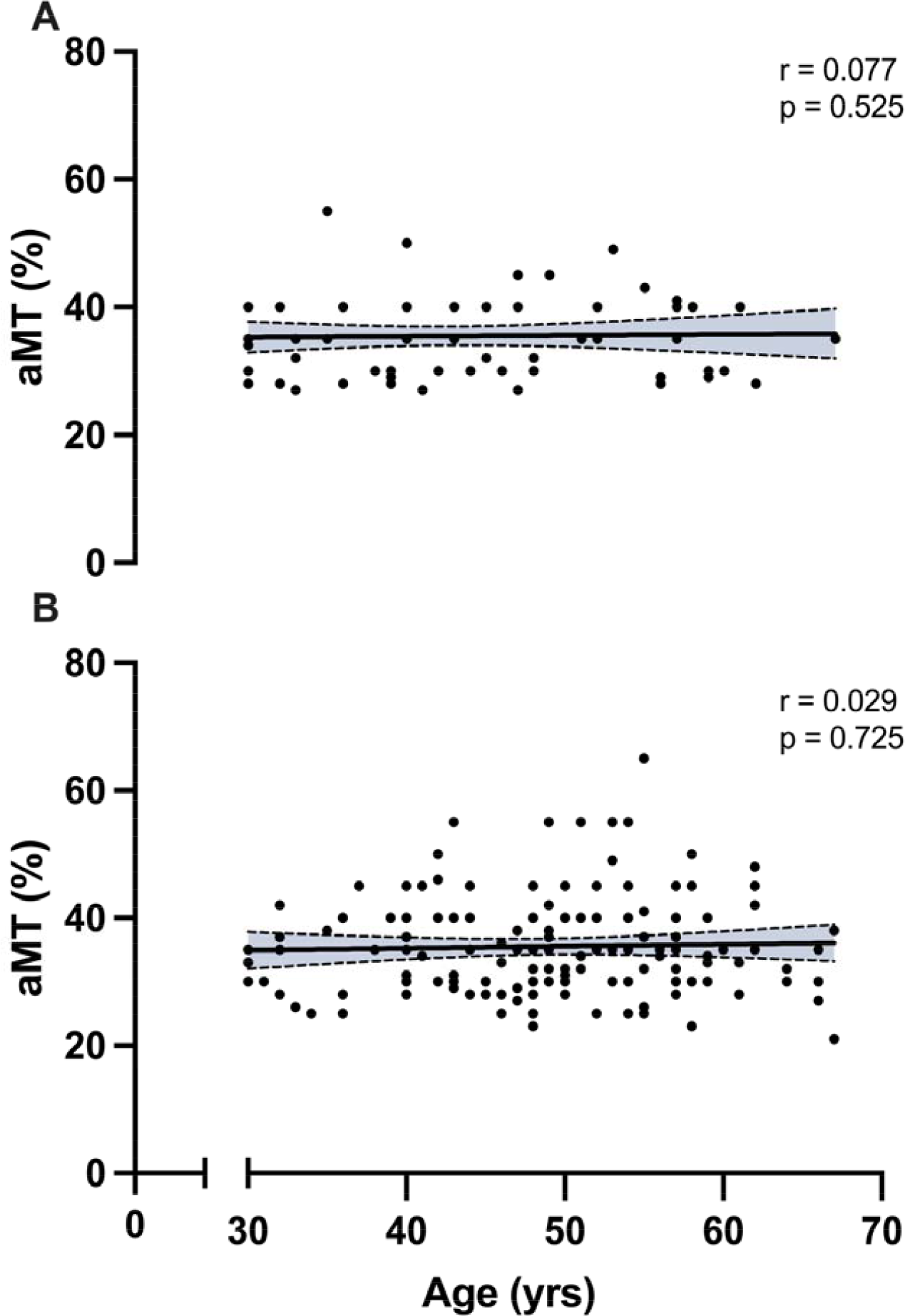
Scatterplots between age and active motor threshold for control (1a) and retired athletes (1b).

**Figure 2.**
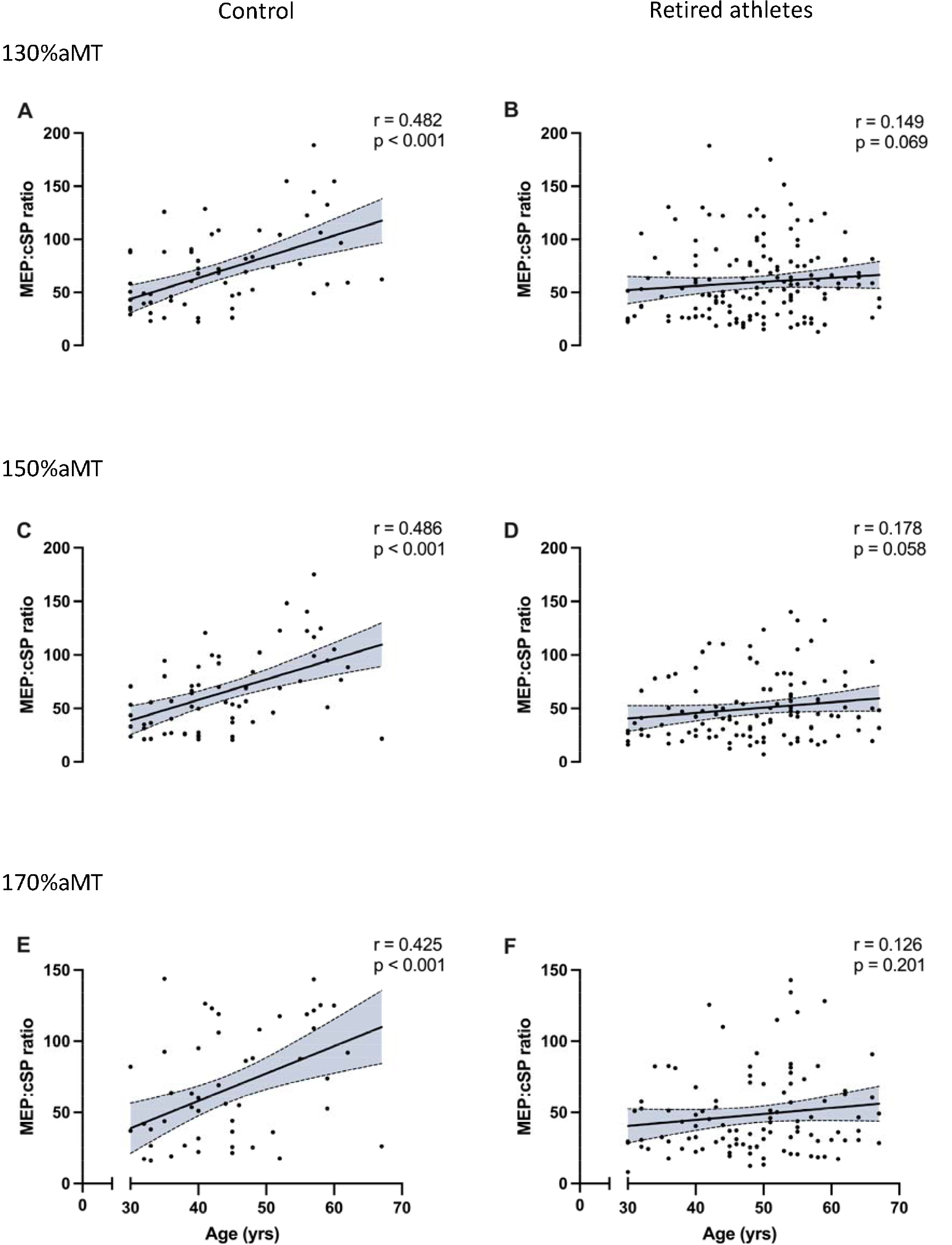
Scatterplots between age and MEP:cSP ratios at 130%, 150% and 170% aMT for controls (2a, 2c, 2e respectively) and retired athletes (2b, 2d, 2f respectively).

SICI showed moderate significant negative correlations for control participants (*rho*=-0.334; *p*=0.01, fig 3a) but were not observed in retired athletes (*rho*=-0.005; *p*=0.956, fig 3b). Similarly, LICI showed moderate significant negative correlations for control participants (*rho*=-0.423; *p*<0.001, fig 3c) but not in retired athletes (*rho*=0.107, *p*=0.243, fig 3d). Comparison between groups showed significant correlations in the control group compared to the retired athlete group for SICI (*Z*=-1.965, *p*=0.049), and LICI (*Z*=-3.734, *p*<0.001),

**Figure 3.**
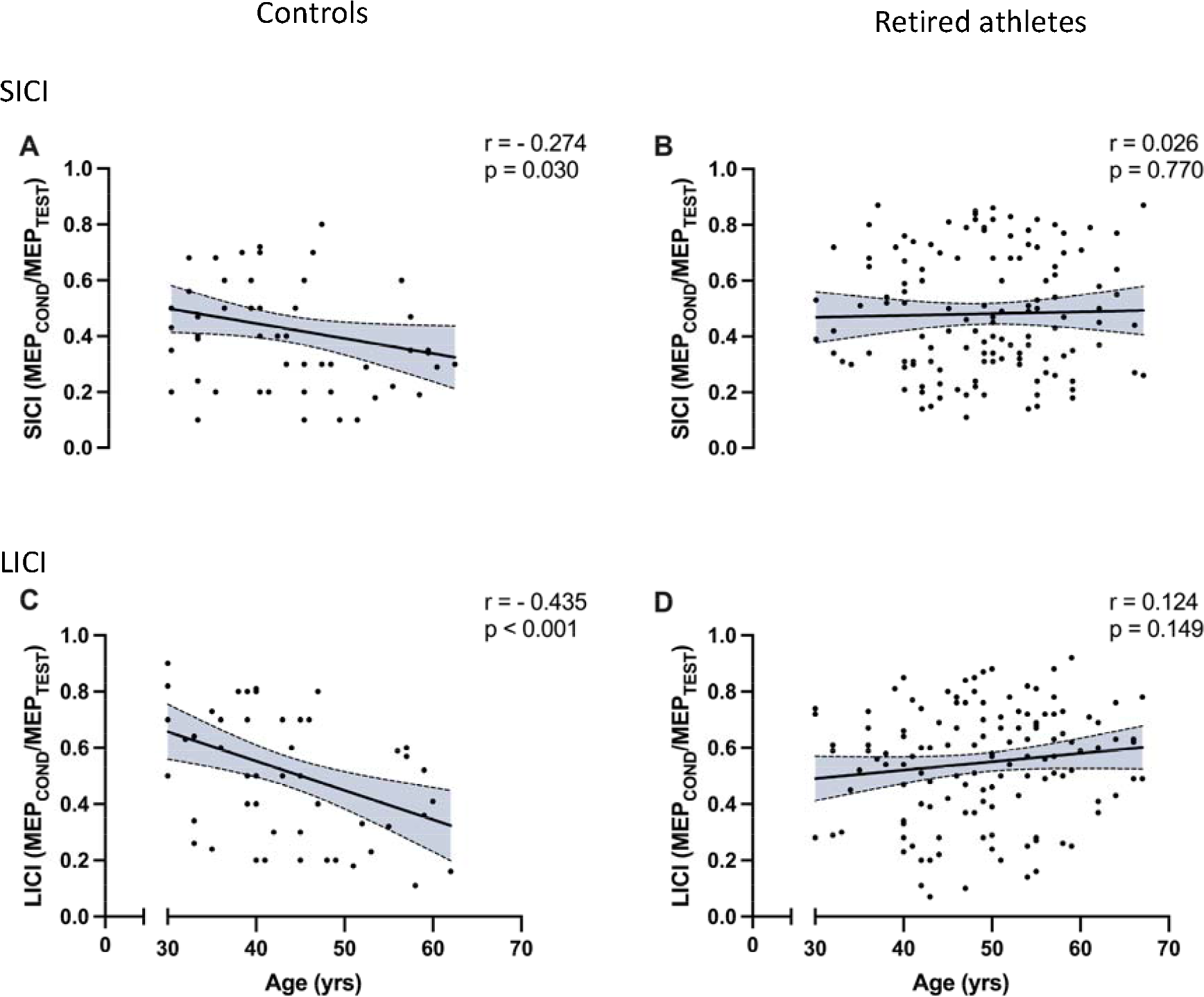
Correlations across age with SICI 3ms (a and b) and LICI 100ms (c and d).

## Discussion

Previous TMS studies have shown inhibitory changes in older retired contact-sport athletes (<50 years) with a history of repetitive neurotrauma [5-8], however TMS studies to date have not included cohorts of retired younger athletes. The aim of this descriptive study was to compare TMS corticomotor and intracortical measures across the lifespan (30-70 years) in those with a history of repetitive head trauma to controls with no history of playing sports. Supporting previous work in healthy ageing [10, 24], we observed increased corticomotor and intracortical inhibition measures in older individuals. When comparing to retired athletes we found significantly stronger correlations of inhibition and age in the control group compared to the retired athlete group, suggesting that across the lifespan retired athletes with progressing age illustrated less inhibition.

The findings in this descriptive study provide further evidence of alterations in GABAergic receptor activity resulting from a history of repetitive head trauma. By what mechanisms repetitive trauma affects GABAergic activity is beyond the scope of this study. However, the data in this study illustrates that retired athletes showing increased inhibition suggests a potential prodromal temporal biomarker as part of overall clinical criteria for traumatic encephalopathy syndrome (TES) [25]. Indeed, TMS evoked potentials have demonstrated the efficacy of prodromal temporal biomarkers in identifying pathophysiology in Alzheimer’s and frontotemporal dementia versus aged-matched controls [26, 27].

Despite this, we acknowledge that intracortical inhibition changes in older individuals are not uniform across studies in both healthy and those with a history of brain injury. As discussed by McGinley et al [10] these differences may be attributed to the target muscle and TMS stimulus protocols. In single pulse TMS, another reason for disparities seen in inhibition changes across studies may also be attributable to cSP duration being measured in isolation and not part of the overall excitation/inhibition balance. Despite being well-known that the intensity of TMS is an influencing confounding factor on MEP and cSP duration [17, 28], whereby increased cSP duration is observed at higher intensities of stimulation until a plateau in duration occurs at high intensities of stimulus [29]. Orth and Rothwell [19], and Škarabot et al [20] have argued that the relationship between stimulus intensity and cSP duration may also be due to the influence of the preceding MEP, which also increases with stimulus intensity until plateau, has on cSP duration [19]. Consequently, the present study sought to present data as cSP:MEP ratio to reflect a balance between excitatory and inhibitory mechanisms. Further to mitigate any influence of differences in motor threshold on changes in cSP duration [29] our study completed a modified stimulus-response curve which also allowed for the comparison of associated MEP amplitude/cSP duration with TMS intensity [29]. Our data showed that with increasing TMS intensities MEP amplitude and cSP duration increased in the control group, but this was not the case in the retired athlete cohort that showed with increasing MEP amplitudes there was not a concurrent lengthening in the cSP suggesting increased net excitability through decreased inhibitory influences [19, 20]. We found similar results in both SICI and LICI ratios suggesting a general change in GABAergic intracortical inhibition.

Our exploratory study is limited by several factors. Firstly, single pulse TMS is a measure of corticomotor physiology and, while it has been argued that cSP represents intracortical networks, we caution on generalising these findings to more global measures of intracortical inhibition. Secondly, SICI and LICI measures were limited to only one ISI (3 ms and 100 ms respectively), and we did not collect intracortical facilitation (ICF). The inclusion of greater ISI has been used to detect intracortical synaptic and cholinergic circuits impairments that have predicted cognitive decline in various dementias [26, 27] and based on the data in this descriptive study, we aim to include greater paired-pulse protocols in future work.

In conclusion, this descriptive study shows the potential of single/paired pulse TMS in differentiating cortiomotor physiology in retired contact athlete cohort compared to an age match controls. We found correlations with increased inhibition associated with progressing age. This data may be useful in supplementing neuropsychological batteries for the assessment of TMS which currently lacks physiological biomarkers.

## Data Availability

All data produced in the present study are available upon reasonable request to the authors

## Acknowledgements

This research received no external funding, nor any support for the work. A.J.P. has previously received partial research funding from the Erasmus+ strategic partnerships program (2019-1-IE01-KA202-051555), Sports Health Check Charity (Australia), Australian Football League, Impact Technologies Inc., and Samsung Corporation, and is remunerated for expert advice to medicolegal practices. A.J.P is also a honorary director of the Concussion Legacy Foundation Austrlaia. No other author has any declaration of interest.

